# Protective effect of influenza vaccine in Brazilian patients hospitalised for severe acute respiratory infection, 2023-2025

**DOI:** 10.64898/2026.09.15.26362995

**Authors:** Emil Kupek

**Affiliations:** Department of Public Health, Centre for Health Sciences, Federal University of Santa Catarina, Brazil

**Keywords:** influenza, mortality, hospitalisation, vaccine, Brazil

## Abstract

**Introduction:** Despite a significant increase in influenza burden in Brazil in 2025, influenza vaccine effectiveness has not included the key end-of-winter period so far. Methods: The key exposure measure was the number of trivalent influenza vaccine doses received within a calendar year. In-hospital mortality and length of stay, use of the intensive care unit (ICU), and ICU stay were outcome measures. Secondary data from January 2003 to October 2025 were retrieved and analysed by Poisson regression.

**Results:** Influenza vaccine uptake among SARI-hospitalised patients dropped from about 70% in 2023 to a 40-50% level in 2024 and 2025. A single vaccine dose reduced outcomes by 10-13% compared to unvaccinated patients. Two doses per year significantly reduced mortality (32%), ICU admissions (52%), and ICU stay (28%).

**Conclusion:** Two doses of influenza vaccine were moderately effective in decreasing in-hospital mortality and stay among SARI-hospitalised influenza patients. Including 50-59-year-olds in a priority vaccination group may significantly reduce influenza-related adverse outcomes.

## Introduction

Various epidemiological reports pointed to a shifting profile of the viruses causing severe acute respiratory infections (SARI) in Brazil during the SARS-CoV-2 pandemic. For example, a nationwide study found a marked reduction in influenza incidence concomitant with a sharp increase in RSV and rhinoviruses during the pandemic in the northeastern state of Bahia ^1^. In the southern state of Rio Grande do Sul in 2023, SARS-CoV-2 was predominant, accounting for 22.92% of SARI, followed by respiratory syncytial virus (RSV) at 15.72% and influenza at 7.59%, of which 71.35% was type A ^2^.

Children in the first year of life were particularly susceptible nationwide, as 2.66% of them had SARI in 2024, with a sharp peak in the second quarter of the year ^3^. Although RSV and rhinoviruses were the most frequent, other viruses of lesser incidence had a higher death rate, e.g. 8.57% for parainfluenza type 4, 2.86% for influenza B, and 2.47% for SARS-CoV-2.

In 2023, influenza mortality decreased in the elderly population, despite a significant reduction in trivalent influenza vaccine coverage to 63% ^4^. However, the number of in-hospital deaths and hospitalisations caused by SARI increased by 157% and 189%, respectively, in 2024 ^5^. Nevertheless, influenza vaccine uptake remained low.

During the pandemic, different influenza strains were observed across Brazil. In the 2021-2023 period, the predominant strain was A/H3N2, which was replaced by A/H1N1pdm09 by the end of the period ^6^.

In the second quarter of 2025, influenza-related mortality in Brazil increased sharply ^7^. However, very few scientific publications related these data to influenza vaccine coverage and key risk factors. The aim of this paper is to present a brief description of the relationship between influenza vaccination, on the one hand, and mortality and hospital stay, on the other, in hospitalised influenza patients.

## Methods

Anonymised individual records of patients hospitalised between January 2023 and July 2025 with Severe Acute Respiratory Syndrome (SARS) were retrieved from a specific registry ^8^, maintained by the Brazilian Ministry of Health. These data also contain information on influenza vaccination, disease outcomes, and risk factors, such as patient age and the presence of various comorbidities. The age groups used for this analysis were 0-4, 5-14, 15-49, 50-64, and 65 years or older.

The influenza case definition included both A and B types, based on laboratory testing, clinical examination, and epidemiological evidence ^8^. Risk factors for influenza included having given birth in the last 45 days, cancer, Down syndrome, asthma, diabetes, immunodepression or immunodeficiency, obesity, chronic cardiovascular, hepatic, neurologic, haematological, renal, and lung disease.

Trivalent influenza vaccine was produced by the “Instituto Butantan” (São Paulo, Brazil) and included the following strains: A/Victoria/4897/2022 (H1N1)pdm09, A/Thailand/8/2022 (H3N2), and B/Austria/1359417/2021 (lineage B/Victoria) ^10^. The vaccination scheme varied according to age and risk factors ^9^, as did its protection effect, whose typical duration was estimated at 12 months ^10^. In the present analysis, the number of influenza vaccine doses within each calendar year between January 2023 and October 2025 was zero, one, and two. Vaccine protection was considered optimal if the recommended vaccination scheme ^9^ was applied, partial if a single vaccine dose was received when the two-dose regimen was in place, and none for unvaccinated individuals.

The outcomes of interest included the number of in-hospital deaths, intensive care unit (ICU) admissions, and duration of hospital/ICU stay.

Descriptive statistics show the vaccine uptake and outcome distributions by age group. Separate univariate Poisson regressions were applied for each outcome, followed by multivariate Poisson regressions which adjusted for age group and calendar year, with the number of influenza hospitalisations as the offset. Vaccine effectiveness was defined as the percentage reduction in incidence ratios of the number of vaccine doses (one or two) received within a calendar year, compared to that of unvaccinated patients, for each outcome.

Statistical uncertainty was expressed by the 95% confidence interval (CI). Stata software was used for data management and statistical analysis ^11^.

## Results

Over the period analysed, 91.48% of influenza cases were laboratory-confirmed, and the remaining diagnoses were based on a combination of clinical, epidemiological and medical imaging criteria. Vaccination status was unknown for 21.18% of the SARS patients eligible to participate in the present study.

Influenza vaccine uptake dropped from about 70% in 2023 to a 40% level in 2024, then recovered about 10% in the following year (Figure 1).

**Figure 1.**
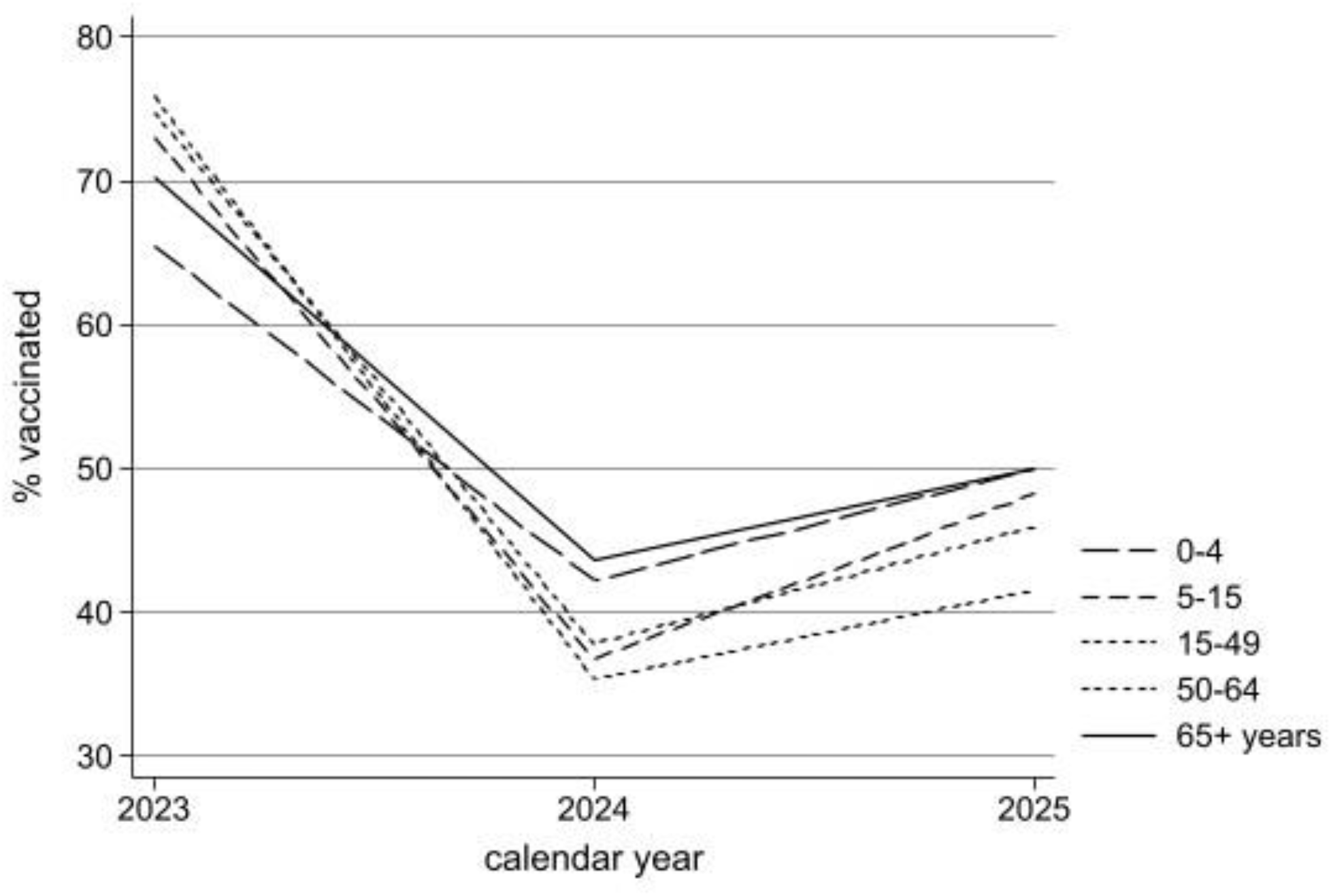
Percentage of trivalent influenza vaccine coverage by age group in Brazil between January 2023 and October 2025

The youngest and oldest age groups had the highest vaccine coverage in 2024 and 2025.

Use of hospital services and in-hospital deaths due to SARI increased from 2024 to 2025 for all age groups except 5-to-14-year-old children and adolescents (Table 1).

**Table 1.**
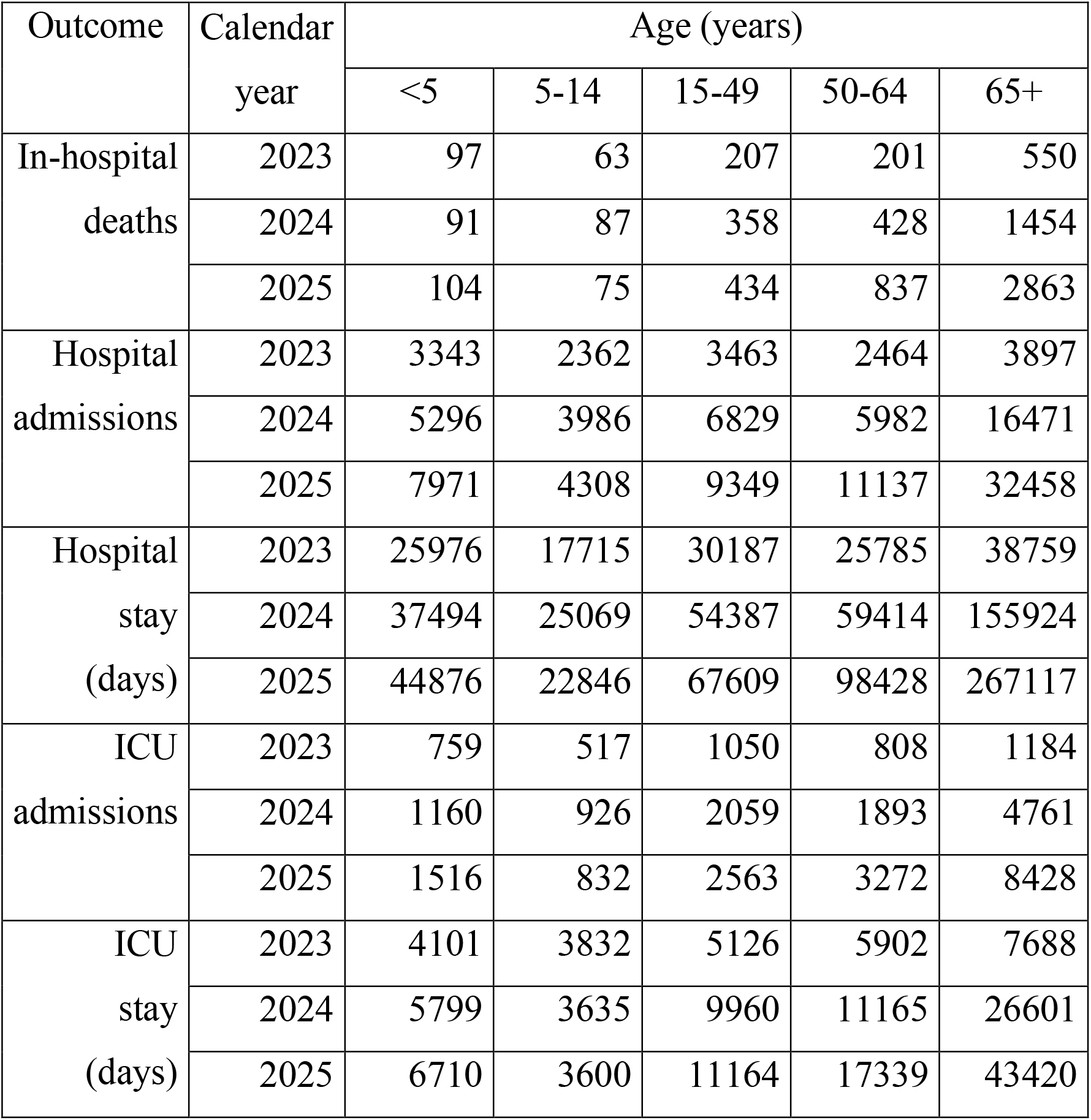
Use of hospital services and in-hospital deaths due to severe acute respiratory infections in Brazil between January 2023 and October 2025.

The largest increase was noted among those 65 years or older.

Univariate Poisson regressions showed more than fourfold increase in hospital mortality and stay among patients 50 years or older compared to those under 5 years (Table 2).

**Table 2.** Separate Poisson regression results for selected outcomes by age group and the number of trivalent vaccine doses received.

| Independent variable | Category | Univariate |  |  |  | Multivariate |  |  |  |
| --- | --- | --- | --- | --- | --- | --- | --- | --- | --- |
|  |  | Mortality | Hospital stay | ICU admission | ICU stay | Mortality | Hospital stay | ICU admission | ICU stay |
| Age (years) | 0-4 | 1.00<sup>a</sup> | 1.00<sup>a</sup> | 1.00<sup>a</sup> | 1.00<sup>a</sup> | 1.00<sup>a</sup> | 1.00<sup>a</sup> | 1.00<sup>a</sup> | 1.00<sup>a</sup> |
|  | 5-14 | 2.89<br>2.54, 3.30 | 2.87<br>2.52, 3.27 | 1.19<br>1.18, 1.20 | 1.18<br>1.17, 1.19 | 1.40<br>1.34, 1.46 | 1.38<br>1.33, 1.44 | 1.34<br>1.31, 1.36 | 1.33<br>1.30, 1.36 |
|  | 15-49 | 1.20<br>1.01, 1.43 | 1.18<br>0.99, 1.40 | 0.94<br>0.94, 0.95 | 0.93<br>0.92, 0.94 | 1.03<br>0.98, 1.09 | 1.02<br>0.97, 1.07 | 1.04<br>1.01, 1.06 | 1.01<br>0.99, 1.04 |
|  | 50-64 | 4.26<br>3.76, 4.83 | 4.35<br>3.84, 4.94 | 1.44<br>1.43, 1.45 | 1.45<br>1.44, 1.47 | 1.47<br>1.41, 1.54 | 1.49<br>1.43, 1.55 | 1.76<br>1.72, 1.79 | 1.81<br>1.78, 1.84 |
|  | 65+ | 5.24<br>4.66, 5.90 | 5.58<br>4.95, 6.29 | 1.34<br>1.33, 1.35 | 1.38<br>1.38, 1.39 | 1.32<br>1.27, 1.37 | 1.35<br>1.30, 1.41 | 1.47<br>1.45, 1.50 | 1.56<br>1.54, 1.59 |
| Vaccine doses received | None | 1.00<sup>a</sup> | 1.00<sup>a</sup> | 1.00<sup>a</sup> | 1.00<sup>a</sup> | 1.00<sup>a</sup> | 1.00<sup>a</sup> | 1.00<sup>a</sup> | 1.00<sup>a</sup> |
|  | One | 0.94<br>0.89, 1.00 | 0.82<br>0.77, 0.87 | 0.92<br>0.91, 0.92 | 0.91<br>0.91, 0.91 | 0.87<br>0.84, 0.90 | 0.88<br>0.85, 0.90 | 0.88<br>0.87, 0.89 | 0.90<br>0.89, 0.91 |
|  | Two | 0.16<br>0.06, 0.42 | 0.57<br>0.21, 1.51 | 0.44<br>0.42, 0.47 | 0.59<br>0.56, 0.62 | 0.68<br>0.54, 0.86 | 0.90<br>0.71, 1.14 | 0.48<br>0.42, 0.54 | 0.72<br>0.64, 0.82 |

<sup>a</sup> reference group

However, the effect was considerably reduced to 47-81% after multivariate adjustment for patient age and calendar year (Table 2). The same adjustment showed that a single dose of vaccine reduced mortality by 13% (10-16%), hospital stay by 12% (10-15%), ICU admissions by 12% (11-13%), and ICU stay by 10% (9-11%), compared to unvaccinated patients. Two doses per year were associated with a 32% (14-46%) lower mortality, a 52% (46-58%) reduction in ICU admissions, and a 28% (18-36%) shorter ICU stay.

## 4. Discussion

This is the first paper to describe the effectiveness of influenza vaccination in reducing in-hospital mortality, hospital/ICU admissions and length of stay, among patients hospitalised for SARI in Brazil between January 2023 and October 2025. Key objectives of the vaccination are to prevent severe outcomes, such as hospitalisation and death, as well as to reduce hospital stay and intensive care services among those hospitalised. While a single vaccine dose was 10-13% effective in reducing in-hospital mortality, hospital/ICU admissions, and stay, two doses enhanced these effects, except for the number of hospital admissions. ICU admissions and stay were 52% and 38% lower, respectively, whereas the 32% mortality reduction was in line with previous studies in Brazil ^9,10^.

Influenza vaccine effectiveness in preventing hospital admission of Brazilian SARI patients was reported at 40.1% (34.8, 45.0%) during the March-September 2025^12^, 30.3% (19.9, 39.4%) in March-July 2024 ^13^, and 31.0 (1.5-51.8%) in 2023 ^14^. In Europe, the vaccine effectiveness ranged from 33% to 56% in the 2024/2025 influenza season ^15^. All these estimates are of a similar magnitude to the present study results.

It is worth noticing that hospitalised patients are the most severely affected among those infected by influenza. Preventing influenza-related hospitalisation is a key vaccination target, which was beyond the scope of the present study. Nevertheless, a sizeable reduction in disease severity among hospitalised influenza patients vaccinated against this infection demonstrated the importance of vaccination for those most susceptible to severe influenza.

Advanced age is associated with increased comorbidity, which in turn increases the risk of severe outcomes. Although 60 years is used to define priority for influenza vaccination in Brazil, the present study results point to including people aged 50-59 years as well. It also highlighted the advantage of two vaccine doses, recommended to occur six months apart ^9^.

The upsurge of influenza-related hospitalisations and deaths in Brazil, after the COVID-19 pandemic was no longer the main health emergency, reached the influenza A(H1N1)_pdm09_2009 level in 2025 ^16^, with an estimated mortality rate of 0.1% in the general population, largely driven by the same strain. The bulk of the mortality burden was observed among older unvaccinated patients with a high number of risk factors; however, the highest hospitalisation rates were registered in unvaccinated children and adolescents. Brazilian health authorities repeatedly appealed to the population to take up the trivalent influenza vaccine at no cost in public primary care facilities. Nevertheless, intensification of vaccine uptake occurred with a significant delay, after the disease toll became more evident in the second quarter of 2025.

Influenza specialists provided a timely warning of the rising burden of influenza in 2024 ^5^ and 2025 ^7^, but vaccine hesitancy was mentioned as a strong obstacle to achieving better vaccine coverage ^5^. Other countries with universal access to the influenza vaccine also found large differences in adherence to the stipulated targets ^17^. In addition to health education, expanding vaccination sites to schools and pharmacies, and customised messages to high-risk groups have been recommended to improve vaccine coverage. Boosting confidence in health authorities, emphasising collective responsibility, and reducing access-to-care constraints may reduce vaccine hesitancy ^18^.

The present study’s strengths include a well-established national surveillance system with a reasonable coverage of key variables of interest. However, the usual limitations of secondary data apply, such as unknown variation in the system’s sensitivity and specificity related to access to medical care and availability of laboratory diagnostic facilities nationwide. A large number of respiratory deaths outside the hospital, classified as “unspecified pneumonia” (J18.9), “unspecified lower respiratory infection” (J22), “unspecified upper respiratory infection” (J06.9), or “respiratory failure” (J96.0/J96.9), may in fact be due to influenza, especially in the northern and northeastern regions of Brazil where underestimation of mortality from chronic diseases was found considerably higher than in other regions ^19, 20^. Also, as a specific contribution of influenza-related sequelae to hospitalisation and mortality from other respiratory infections is notoriously difficult to disentangle, a broad definition of the influenza burden should be assumed here. Finally, although individual-level data were used to estimate vaccine effectiveness in a hospital setting, these do not differentiate between individual and herd immunity, nor do they account for inaccuracy in self-reported number of influenza vaccine doses received.

The choice of in-hospital deaths as a plausible lower bound of the overall influenza mortality was driven by the fact that this was the only real-time data available, because the Brazilian Ministry of Health web report on mortality by specific causes usually takes a year-long delay, thus making this information of little practical use to modify an ongoing immunisation campaign. Consequently, adding out-of-hospital influenza deaths would provide a better estimate of the overall toll of this disease.

## Conclusions

Compared to unvaccinated patients hospitalised for influenza, those who received two doses of trivalent vaccine per year had a 32% lower mortality, a 52% reduction in ICU admissions, and a 28% shorter ICU stay. Including 50-59-year-olds in a priority vaccination group may significantly reduce influenza-related adverse outcomes.

## Data Availability

https://opendatasus.saude.gov.br/dataset/srag-2021-a-2024

